# Ergonomics knowledge, attitude, and practice among biomedical scientists

**DOI:** 10.1101/2021.07.25.21261088

**Authors:** Nasar Alwahaibi, Ibrahim Al Abri, Mallak Al Sadairi, Samira Al Rawahi

**Affiliations:** Department of Allied Health Sciences, College of Medicine and Health Sciences, Sultan Qaboos University. Oman; Department of Pathology, Sultan Qaboos University Hospital, Sultan Qaboos University. Oman

**Keywords:** Attitude, Biomedical scientists, Ergonomics, Knowledge, Practice

## Abstract

Biomedical scientists (BMSs) are important professionals for healthcare services as they help in the detection, diagnosis, and treatment of numerous diseases. To the best of our knowledge, this is the first study to assess ergonomics knowledge, attitude, and practice among BMSs. A cross-sectional study was carried out among BMSs using a self-constructed questionnaire. The association between these parameters and various risk factors was measured using the Chi-square test. The study included 110 BMSs. Females represented 68.2% and 45.5% were in the age of 25-34. Good ergonomics knowledge showed in 54.5% and 82.7% showed high positive attitude. However, poor ergonomics showed in 83.5%. There was no significant interrelation between the three parameters. A significant association was found between the male gender (0.040), more than 20 working experience (0.016), and good ergonomics practice. Biomedical scientists have good knowledge, high attitude but the poor practice of ergonomics. Ergonomics training and practice should be firmly enhanced among these healthcare professionals.

## Introduction

Biomedical scientists (BMSs) are important professionals for healthcare services as they help in the detection, diagnosis, and treatment of numerous diseases. However, in day-to-day practice, BMSs are continuously exposed to various hazards such as chemical, physical, mechanical, electrical, and biological, as well as they are prone to infectious specimens (1). The most common daily activities for BMSs include pipetting, microscopy, and work on standing positions. With the time, this may affect body organs such as muscles, joints, tendons, nerves, ligaments, cartilage, and spinal discs (2). These musculoskeletal disorders (MSDs) can affect the efficiency and speed of the work and eventually lead to a disruption in the healthcare system (3). It has been reported that MSDs are one of the common health problems among all health professionals (4).

The goal of laboratory ergonomics is to reduce as much as possible the MSDs among BMSs by providing safe and comfortable working environment. Due to the nature of their work in the medical laboratory, BMSs are at risk for many health problems. Previous studies have focused mainly on the dentistry as an important profession in the healthcare system. However, scanty studies have evaluated the knowledge, attitude, and practice of ergonomics among BMSs. Thus, this study aimed to assess those ergonomics parameters among biomedical scientists.

## Methods

This is a cross-sectional observational study that was conducted in the year of 2020 at the Sultan Qaboos University Hospital (SQUH) and Sultan Qaboos University (SQU). The study is approved from the Medical Research Ethics Committee, College of Medicine and Health Sciences, SQU (SQU-EC/095/2020). Preceding the study, a detailed procedure of the study was explained, a voluntary consent form was also signed by each participant before the study.

The sample size was calculated using the infinite population sample size formula: n = {N × Z^2^ × p × (1–p)} / {d^2^ × (N–1) + Z^2^× p × (1 – p) where N : Population size (The total number of BMS working at SQU and SQUH) = 170, Z (Standard value with confidence level 95%) =1.96, d(Permissible error on either side) = 10%, p (Proportion of the characteristic under the study). At the present, there is no available information on the proportion of MSDs among BMS. Therefore, the *P*-value was obtained from a pilot study that was conducted among 20 participants who fulfilled the research criteria. Those who participated in the pilot study were excluded from the study. The *P*-value was found equal to 63.5%. The sample size was calculated as 97. After sampling, the number was increased 15% to avoid non-response or inappropriately filled questions and that number was randomly selected from the overall population. The reliability of the questionnaire was calculated by Cronbach alpha in the pilot study and found to be 0.675.

A self-constructed questionnaire was used to obtain knowledge, attitude, and practice among biomedical scientists. It was designed utilizing literature review and other questionnaires of other populations (5-6).

The questionnaire contained a brief description of the study. It was designed to contain a mix of positive and negative questions to avoid the false-positive results because of the inappropriately filled questions.

The questionnaire used in this study consisted of four sections. The first section was socio-demographic characteristics such as age, sex, marital status, physical exercise, heavy work at home, nationalities, qualifications, number of working years, specialities, BMS grade, working hours, work shifts and overtime. The second section consisted of eight questions related to the ergonomics knowledge, such as meaning, benefits, and principles of ergonomics. The third section consisted of eight questions related to the ergonomics attitude such as ergonomics education, distribution of the work, and adjusting the workplace. The fourth section consisted of nine questions related to the ergonomics practice such as wearing comfortable shoes, using comfortable positions, and relaxing eyes and neck.

## Statistical analysis

For the three categories: Yes, some extent, and no, the scores were two, one and zero, respectively. The total score of knowledge ranged from zero to 16. Achievement of more than 66.66% of the maximum score considered as good knowledge. Attitude was assessed by strongly agree, agree, neutral, disagree, or strongly disagree. The score of each question ranged from 1–5 giving a total score range of 5 – 40. Those who achieved more than 66.66% regarded as having a positive attitude. Practice was evaluated like the knowledge. The total score ranged from zero to 18. Achievement of more than 66.66% of the maximum score considered as good practice.

The data were analyzed using Statistical Package for Social Science (SPSS) version 25 software (SPSS Inc., Chicago, USA). Frequencies and percentages were used to represent the categorical data such as age, gender, and specialty. Continuous data were presented as mean and standard deviation. A Chi-square test was performed to measure the significant association between risk factors and level of knowledge, attitude, and practice. The *P*-value was considered significant if it was equal to or less than 0.05.

## Results

A total of 110 BMSs were included in this study after excluding those who did not fit the inclusion criteria. The majority were females (68.2%) and 73.6% were married. The majority of the BMSs (45.5%) were in the age group of 25-34. Surprisingly, the number of those who were doing regular physical exercises was nearly equal to those who were not with 50.9% and 49.1%, respectively (Table 1). 30.0% of BMSs reported to have heard of the term “ergonomics, 37.3% knew the benefits of ergonomics application, and 47.3% knew the health hazard of their work without ergonomics (Table 2).

**Table 1:** Sociodemographic characteristics among biomedical scientists

| Characteristic | Number (110) | Percent (%) |
| --- | --- | --- |
| Gender |  |  |
| • Males | 35 | 31.8 |
| • Females | 75 | 68.2 |
| Age group |  |  |
| • <25 | 1 | 0.9 |
| • 25-34 | 50 | 45.5 |
| • 35-44 | 42 | 38.2 |
| • 45-54 | 11 | 10.0 |
| • >54 | 6 | 5.5 |
| Marital status |  |  |
| • Single | 29 | 26.4 |
| • Married | 81 | 73.6 |
| Nationality |  |  |
| • Omani | 89 | 80.9 |
| • Non-Omani | 21 | 19.1 |
| Off work physical activity |  |  |
| • Yes | 56 | 50.9 |
| • No | 54 | 49.1 |
| Specialty |  |  |
| • Hematology | 30 | 27.3 |
| • Biochemistry | 23 | 20.9 |
| • Histopathology | 21 | 19.1 |
| • Microbiology and Immunology | 20 | 18.2 |
| • Genetics | 8 | 7.3 |
| • Anatomy | 6 | 5.5 |
| • Physiology | 2 | 1.8 |
| Qualification |  |  |
| • Diploma | 7 | 6.4 |
| • Bachelor | 71 | 64.5 |
| • Master | 28 | 25.5 |
| • PhD | 4 | 3.6 |
| Designation |  |  |
| • Junior BMSs | 53 | 48.2 |
| • Senior BMSs | 38 | 34.6 |
| • Chief BMSs | 14 | 12.7 |
| • Superintendent | 3 | 2.7 |
| • Researcher | 2 | 1.8 |
| Total work years as BMSs |  |  |
| • <20 | 90 | 81.8 |
| • ≥20 | 20 | 18.2 |

**Table 2:**
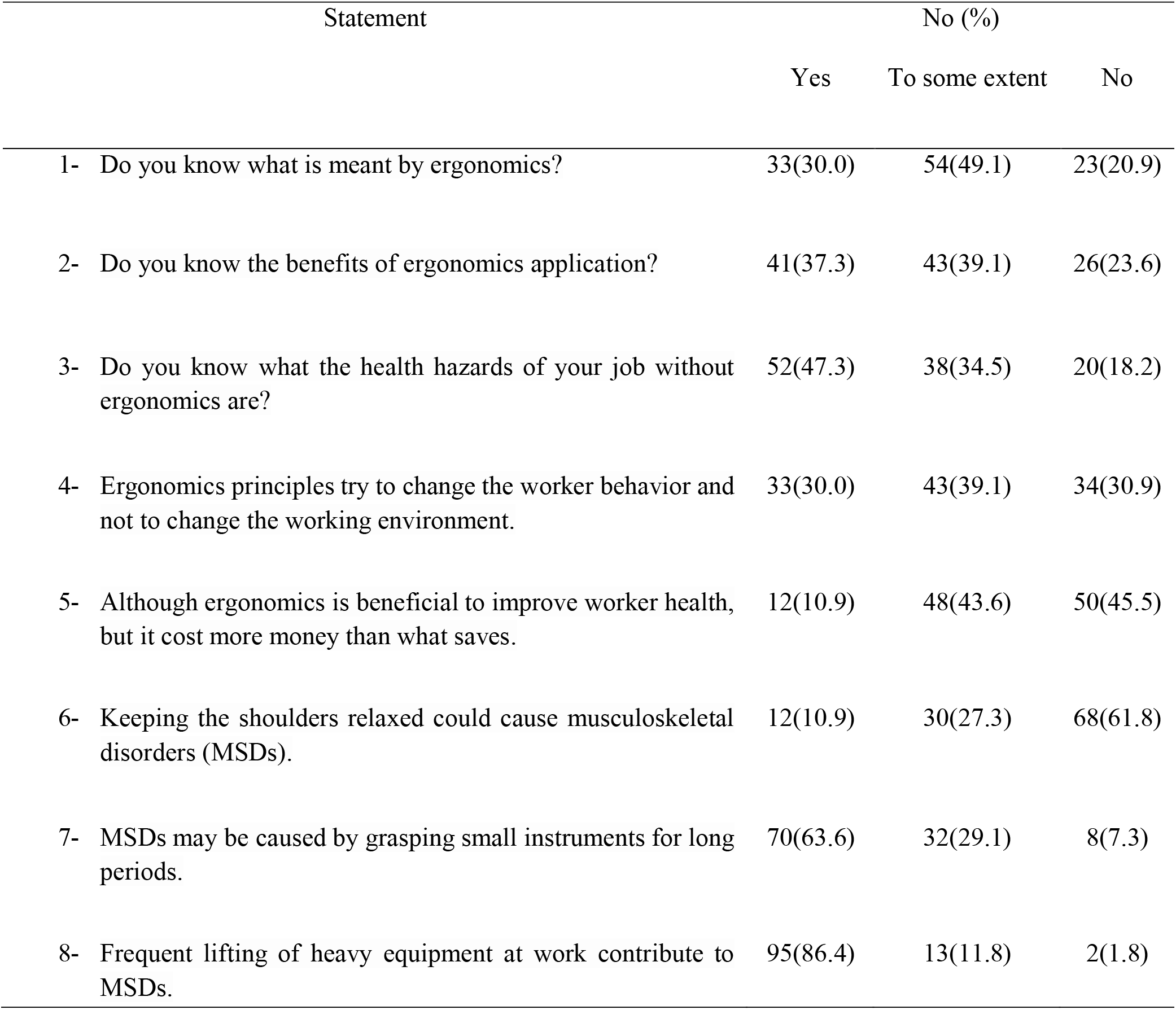
Ergonomics knowledge among biomedical scientists

| Statement | No (%) |  |  |
| --- | --- | --- | --- |
|  | Yes | To some extent | No |
| 1- Do you know what is meant by ergonomics? | 33(30.0) | 54(49.1) | 23(20.9) |
| 2- Do you know the benefits of ergonomics application? | 41(37.3) | 43(39.1) | 26(23.6) |
| 3- Do you know what the health hazards of your job without ergonomics are? | 52(47.3) | 38(34.5) | 20(18.2) |
| 4- Ergonomics principles try to change the worker behavior and not to change the working environment. | 33(30.0) | 43(39.1) | 34(30.9) |
| 5- Although ergonomics is beneficial to improve worker health, but it cost more money than what saves. | 12(10.9) | 48(43.6) | 50(45.5) |
| 6- Keeping the shoulders relaxed could cause musculoskeletal disorders (MSDs). | 12(10.9) | 30(27.3) | 68(61.8) |
| 7- MSDs may be caused by grasping small instruments for long periods. | 70(63.6) | 32(29.1) | 8(7.3) |
| 8- Frequent lifting of heavy equipment at work contribute to MSDs. | 95(86.4) | 13(11.8) | 2(1.8) |

Regarding the attitude, 59.1% of the BMSs strongly agreed that ergonomics education must be part of the biomedical curriculum, and 68.2% strongly agreed that distributing the work equally between the workers makes it easier. However, 20.0% of them strongly disagreed preferring to bend their head forward instead of adjusting workspace for better viewing (Table 3).

**Table 3:** Ergonomics attitude among biomedical scientists

| Statement | No (%) |  |  |  |  |
| --- | --- | --- | --- | --- | --- |
|  | Strongly agree | Agree | Neutral | Disagree | Strongly disagree |
| 1- Ergonomics education must be a part of the biomedical curriculum. | 65(59.1) | 35(31.8) | 9(8.2) | 0(0.0) | 1(0.9) |
| 2- It is important to distribute the work equally between us because it makes the work easier. | 75(68.2) | 33(30.0) | 1(0.9) | 0(0.0) | 1(0.9) |
| 3- I prefer to bend my head forward instead of adjusting the workspace for better viewing. | 6(5.5) | 18(16.3) | 24(21.8) | 40(36.4) | 22(20.0) |
| 4- Always I should bend my back while working because it makes my work easier. | 9(8.2) | 31(28.2) | 24(21.8) | 27(24.5) | 19(17.3) |
| 5- To finish my work on time, I prefer to attain the same position (e.g., Sitting) for long periods while working instead of changing my posture. | 11(10.0) | 26(23.6) | 18(16.4) | 39(35.5) | 16(14.5) |
| 6- Forceful hand movements while working enables me get work done on time regardless of the consequences that may occur. | 3(2.7) | 35(31.8) | 23(20.9) | 34(30.9) | 15(13.6) |
| 7- Doing exercises like stretching, walking etc. are important to be more productive at work. | 66(60.0) | 38(34.5) | 6(5.5) | 0(0.0) | 0(0.0) |
| 8- I should always try to avoid putting my angled elbow in direct contact with the work surface for a long period. | 26(23.6) | 47(42.7) | 28(25.5) | 7(6.4) | 2(1.8) |

Regarding the practice of BMSs, 72.8% wore comfortable shoes while standing at work, 46.8% found enough space to put their legs and feet comfortably, and 45.0% were trying to avoid pressure on their hands and arms from sharp edges (Table 4).

**Table 4:** Ergonomics practice among biomedical scientists

|  | Statement | No (%) |  |  |
| --- | --- | --- | --- | --- |
|  |  | Yes | Some-time | No |
| 1- | When you are working at the standing position, do you wear comfortable shoes? | 79(72.8) | 23(21.1) | 7(6.4) |
| 2- | Do you find enough space to put your legs and feet in a comfortable position? | 51(46.8) | 38(34.9) | 20(18.3) |
| 3- | Are your hands or arms subjected to pressure from sharp edges on work surfaces? | 22(20.2) | 38(34.8) | 49(45.0) |
| 4- | Do all task requirements are visible from comfortable positions? | 33(30.3) | 53(48.6) | 23(21.1) |
| 5- | Do you avoid raising your arm above your elbow level while you are working? | 20(18.3) | 49(45.0) | 40(36.7) |
| 6- | Do you avoid twisting or bending your wrist? | 29(26.6) | 47(43.1) | 33(30.3) |
| 7- | Do you stop after at least 15 minutes for a moment to relax your eyes and neck? | 7(6.4) | 52(47.7) | 50(45.9) |
| 8- | Do you stop every 30-60 minutes to get up to stretch and move? | 12(11.0) | 57(52.3) | 40(36.7) |
| 9- | While you are holding objects (e.g., Forceps), do you alternate between your fingers holding them? (ex. Thumb and index or index and middle finger) | 6(5.5) | 32(29.4) | 71(65.1) |

There was no significant association between sociodemographic factors and ergonomics knowledge and attitude. However, male gender and more than 20 years work experience showed a significant association with good ergonomics practice (Table 5).

**Table 5:** Ergonomics knowledge, attitude, and practice of biomedical scientists and associations with their sociodemographic characteristics

| Sociodemographic characteristics | Knowledge<br>No (%) |  | *P*<br>value | Attitude<br>No (%) |  | *P*<br>value | Practice<br>No (%) |  | *P*<br>value |
| --- | --- | --- | --- | --- | --- | --- | --- | --- | --- |
|  | Good | Bad |  | Positive | Negative |  | Good | Poor |  |
| Gender |  |  |  |  |  |  |  |  |  |
| • Male | 21(60.0) | 14(40.0) | 0.562 | 28(80.0) | 7(20.0) | 0.806 | 10(28.6) | 25(71.4) | 0.040 |
| • Female | 39(52.0) | 36(48.0) |  | 63(84.0) | 12(16.0) |  | 8(10.8) | 66(89.2) |  |
| Age group |  |  |  |  |  |  |  |  |  |
| • <25 | 0(0.0) | 1(100.0) | 0.678 | 1(100.0) | 0(0.0) | 0.813 | 1(100.0) | 0(0.0) | 0.018* |
| • 25-34 | 28(56.0) | 22(44.0) |  | 41(82.0) | 9(18.0) |  | 5(10.0) | 45(90.0) |  |
| • 35-44 | 21(50.0) | 21(50.0) |  | 34(81.0) | 8(19.0) |  | 6(14.3) | 36(85.7) |  |
| • 45-54 | 7(63.6) | 4(36.4) |  | 9(81.8) | 2(18.2) |  | 4(40.0) | 6(60.0) |  |
| • >54 | 4(66.7) | 2(33.3) |  | 6(100.0) | 0(0.0) |  | 2(33.3) | 4(66.7) |  |
| Marital status |  |  |  |  |  |  |  |  |  |
| • Single | 15(51.7) | 14(48.3) | 0.890 | 23(79.3) | 6(20.7) | 0.779 | 7(24.1) | 22(75.9) | 0.318 |
| • Married | 45(55.6) | 36(44.4) |  | 68(84.0) | 13(16.0) |  | 11(13.8) | 69(86.3) |  |
| Qualification |  |  |  |  |  |  |  |  |  |
| • Diploma | 5(71.4) | 2(28.6) | 0.627 | 6(85.7) | 1(14.3) | 0.531 | 3(50.0) | 3(50.0) | 0.120 |
| • Bachelor | 38(53.5) | 33(46.5) |  | 60(84.5) | 11(15.5) |  | 11(15.5) | 60(84.5) |  |
| • Master | 14(50.0) | 14(50.0) |  | 21(75.0) | 7(25.0) |  | 3(10.7) | 25(89.3) |  |
| • PhD | 3(75.0) | 1(25.0) |  | 4(100.0) | 0(0.0) |  | 1(25.0) | 3(75.0) |  |
| Designation |  |  |  |  |  |  |  |  |  |
| • Researcher | 1(50.0) | 1(50.0) | 0.509 | 2(100.0) | 0(0.0) | 0.878 | 0(0.0) | 2(100.0) | 0.702 |
| • Junior BMSs | 29(54.7) | 24(45.3) |  | 43(81.1) | 10(18.9) |  | 7(13.2) | 46(86.8) |  |
| • Senior BMSs | 21(55.3) | 17(44.7) |  | 32(84.2) | 6(15.8) |  | 8(21.6) | 29(78.4) |  |
| • Chief BMSs | 6(42.9) | 8(57.1) |  | 12(85.7) | 2(14.3) |  | 2(14.3) | 12(85.7) |  |
| • Superintendent | 3(100.0) | 0(0.0) |  | 2(66.7) | 1(33.3) |  | 1(33.3) | 2(66.7) |  |
| Total work years as BMSs |  |  |  |  |  |  |  |  |  |
| • <20 | 47(52.2) | 43(47.8) | 0.430 | 74(82.2) | 16(17.8) | 1.000 | 11(12.2) | 79(87.8) | 0.016 |
| • ≥20 | 13(65.0) | 7(35.0) |  | 17(85.0) | 3(15.0) |  | 7(36.8) | 12(63.2) |  |

\* This cannot be considered as significant even though *P* value is less than 0.05 because more than 20% of cells have expected count less than 5 and the minimum expected count is less than 1 so, the chi square test cannot be used.

Overall, BMSs showed good knowledge and attitude in 54.5%, 82.7%, respectively. However, good practice was seen in only 16.5% (Figure 1). 86.7% of the BMSs with good knowledge have a positive attitude but without any significant association between knowledge and attitude. Good practice was seen in 18.6% of those who have good knowledge with no relationship between knowledge and practice. In addition, there was no relationship detected between attitude and practice (Table 6).

**Figure 1:**
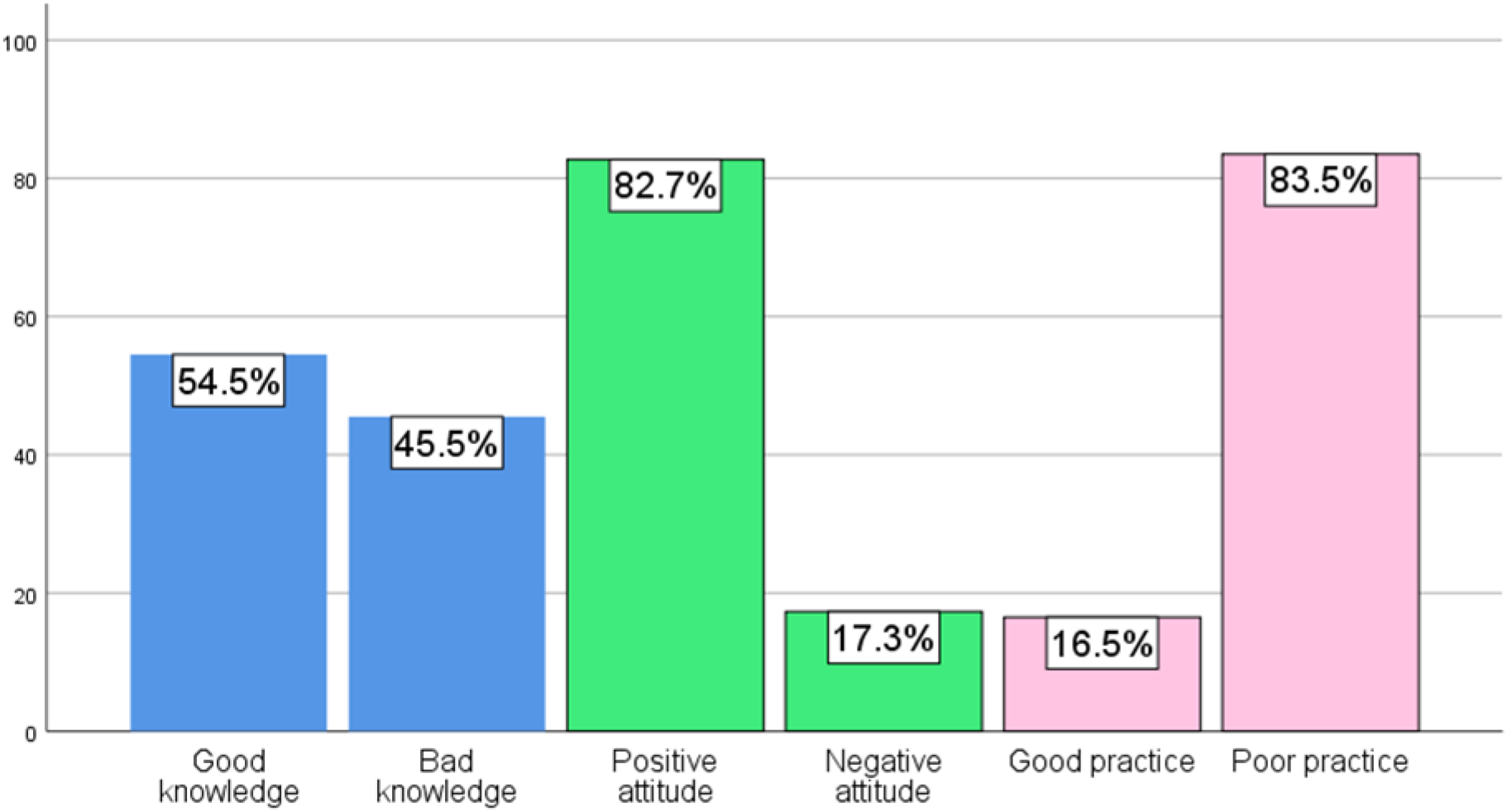
Level of knowledge, attitude, and practice among biomedical scientists

**Table 6:** The relationship between knowledge, attitude, and practice

|  | Attitude |  | *P* value | Practice |  | *P* value |
| --- | --- | --- | --- | --- | --- | --- |
|  | Positive<br>No (%) | Negative<br>No (%) |  | Good<br>No (%) | Poor<br>No (%) |  |
| Knowledge |  |  |  |  |  |  |
| Good | 52(86.7) | 8(13.3) | 0.345 | 11(18.6) | 48(81.4) | 0.695 |
| Bad | 39(78.0) | 11(22.0) |  | 7(14.0) | 43(86.0) |  |
| Attitude |  |  |  |  |  |  |
| Positive |  |  |  | 14(15.6) | 76(84.4) | 0.514 |
| Negative |  |  |  | 4(21.1) | 15(78.9) |  |

## Discussion

The use of laboratory investigation is very important as 70% of all medical decisions are affected by the results of laboratory analysis (7-9). The application of ergonomics guidelines in the field of medical laboratory helps to minimize MSDs. MSDs are one of the common health problems among all health professionals. Several studies reported high levels of MSDs among medical laboratory scientists (10-14). These high MSDs among BMSs will eventually affect the healthcare service. Health, productivity, and well-being are important characters for BMSs (15). It is expected that good knowledge, positive attitude, and good practice of ergonomics would minimize MSDs among BMSs.

To the best of our knowledge, this is the first study to assess ergonomics knowledge, attitude, and practice among BMSs. In the literature review, we found only one study evaluated the ergonomics knowledge only among medical laboratory scientists in Nigeria (15). It is important to note that several instruments and products have been modified in the clinical laboratory to minimize MSDs. For example, laboratory chairs are made with comfort to adjust height, back, and legs. In addition, BMSs while purchasing any instrument, keep in their mind, the ergonomics part of these machines. Although that 93.6% of the BMSs have high qualifications, the present study showed that only 54.5% of BMSs had a good ergonomics knowledge. This finding is higher than other study that reported 25.5% (27 of 106) knowledge of ergonomics among medical laboratory scientists in Nigeria (15). We did not observe any statistically significant association between ergonomics knowledge and gender, age, marital status, qualifications, designation, specialty, and work experience. In comparison with the study in Nigeria, they showed that ergonomics knowledge was significantly associated with male gender. Other risk factors such as qualifications, affiliations, work experience, and specialty did not affect the ergonomics knowledge (15).

The current study showed very high positive ergonomics attitude (82.7%) among BMSs. However, gender, age, marital status, qualifications, designation, specialty, and work experience did not statistically affect the ergonomics attitude. When we asked BMSs if ergonomics education must be a part of the biomedical curriculum, 90.9% (100 of 110) agreed with this concept. In addition, 98.2% (108 of 110) believed that it is important to distribute the work equally between colleagues as it makes the work easier.

Surprisingly, good ergonomics practice was noticed in only 16.5% among BMSs. In addition, there was a statistically significant association between male gender and good practice. This association could be related to the fact that males are less stress than females. Thus, males can do their job more ergonomically. Furthermore, there was a significant association between work experience (more than 20 years) and good practice. Those with more work experience have adopted well to the occupational safety procedure in the medical laboratory.

Previously, we reported that the overall prevalence of laboratory–related musculoskeletal disorders (LMSDs) among BMSs in the last 12 months in any parts of the body was 77.1%. Neck complaint was the highest, followed by shoulders, lower back, upper back, knees, ankle or feet, wrists or hands, elbows, and thighs or hip, with 50.6%, 49.4%, 43.4%, 34.9%, 30.1%, 26.5%, 24.1 %, 9.6%, 10.8%, respectively. In addition, 65.57% of BMSs had irregular symptoms of LMSDs, 54.10% experienced moderate pain due to these symptoms, and 44.26% had symptoms that persisted from hours to days (16). Thus, poor ergonomics practice of BMSs resulted in high levels of LMSDs.

Despite that, BMSs had a reasonably good ergonomics knowledge, high positive attitude, and poor practice, we did not observe any statistically significant association between knowledge and attitude, knowledge and practice, or attitude and practice. Thus, BMSs have the ergonomics knowledge and attitude but did not result in a good daily practice. In the medical laboratory, good ergonomics practice leads to less MSDs. In fact, good knowledge, attitude, and practice of ergonomics would help BMSs in taking sufficient precautions during their laboratory work as well as adjusting their working environment accordingly. The findings of this study strongly recommend the use of laboratory ergonomics checklist that would assess BMSs at their workplace and subsequently minimize LMSDs.

This study has some limitations. First, due to the COVID-19 pandemic, we were not able to spread the questionnaire face to face to the BMSs. Second, the study was restricted to BMSs from a single hospital and university, even though this hospital serves as a tertiary referral hospital in Oman and the university is the national university of the Sultanate of Oman. Third, ergonomics practice was measured by questionnaire only and not by observation. Finally, the lack of similar and relevant studies for a better comparison.

In conclusion, biomedical scientists have a good knowledge, high attitude but poor practice of ergonomics. Ergonomics training and practice should be firmly enhanced among these healthcare professionals.

## Data Availability

All data referred to this manuscript are available.

## Acknowledgments

The authors would like to thank all BMSs who participated in this study.

## References

1. Jegathesan M, Chin CS, Lim HH. Safety in pathology laboratories. Malays J Pathol 1988;10:1–5.

2. Haile EL, Taye B, Hussen F. Ergonomic workstations and work-related musculoskeletal disorders in the clinical laboratory. Lab Med 2012;43:11–19.

3. Dong H, Zhang Q, Liu G, Shao T, Xu Y. Prevalence and associated factors of musculoskeletal disorders among Chinese healthcare professionals working in tertiary hospitals: A cross-sectional study. BMC Musculoskelet Disord 2019;20(175):1–8.

4. Tinubu BM, Mbada CE, Oyeyemi AL, Fabunmi AA. Work-related musculoskeletal disorders among nurses in Ibadan, south-west Nigeria: a cross-sectional survey. BMC Musculoskelet Disord 2010;11:12.

5. El-Sallamy RM, Atlam SA, Kabbash I, El-Fatah SA, El-Flaky A. Knowledge, attitude, and practice towards ergonomics among undergraduates of Faculty of Dentistry, Tanta University, Egypt. Environ Sci Pollut Res Int 2018;25(31):30793–30801.

6. Ephraim-Emmanuel B, Ogbomade R, Idumesaro B, Ugwoke I. Knowledge, attitude and practice of preventing the occurrence of work-related musculoskeletal disorders among doctors in University of Port-Harcourt Teaching Hospital. J Med Res Innov 2019;3(2):e000161.

7. Forsman RW. Why is the laboratory an afterthought for managed care organizations? Clin Chem 1996;42:813–816.

8. Hallworth MJ. The ‘70% claim’: what is the evidence base? Ann Clin Biochem 2011;48:487–488.

9. Rohr UP, Binder C, Dieterle T, Giusti F, Messina CG, Toerien E, et al. The value of in vitro diagnostic testing in medical practice: a status report. PLoS One. 2016;11:e0149856.

10. Sadeghian F, Kasaeian A, Noroozi P, Vatani J, Taiebi SH. Psychosocial and individual characteristics and musculoskeletal complaints among clinical laboratory workers. Int J Occup Saf Ergon 2014;20:355–361.

11. AlNekhilan AF, AlTamimi AM, AlAqeel BY, AlHawery AA, AlFadhel SF, Masuadi EM. Work-related musculoskeletal disorders among clinical laboratory workers. Avicenna J Med 2020;23:10(1):29–34.

12. Maulik S, Iqbal R. Occupational health and musculoskeletal symptoms among Indian medical laboratory technicians. JOHE 2013;2:82–92.

13. Ramadan PA, Ferreira MJ. Risk factors associated with the reporting of musculoskeletal symptoms in workers at a laboratory of clinical pathology. Ann Occup Hyg 2006;50(3):297–303.

14. Maulik S, Iqbal R, De A, Chandra AM. Evaluation of the working posture and prevalence of musculoskeletal symptoms among medical laboratory technicians. J Back Musculoskelet Rehabil 2014;27(4):453–461.

15. Oladeinde B, Ekejindu I, Omoregie R, Aguh O. Awareness and knowledge of ergonomics among Medical Laboratory Scientists in Nigeria. Ann Med Health Sci Res 2015;5:423–427.

16. Alwahaibi N, Al Sadairi M, Al Abri I, Al Rawahi S. Prevalence of laboratory-related musculoskeletal disorders among biomedical scientists. medRxiv 2021.06.04.21258372; doi:https://doi.org/10.1101/2021.06.04.21258372

